# Trends, wealth inequalities and the role of the private sector in caesarean section in the Middle East and North Africa: a repeat cross-sectional analysis of population-based surveys

**DOI:** 10.1101/2021.04.14.21255453

**Authors:** Stephen McCall, Aline T. Semaan, Noon Altijani, Charles Opondo, Mohamed Abdel-fattah, Tamar Kabakian-Khasholian

## Abstract

**Objective:** To examine trends and variations of caesarean section by economic status and type of healthcare facility in Arab countries in Middle East and North Africa (MENA).

**Methods:** Secondary data analysis of nationally representative household surveys across nine Arab countries in MENA. The study population was women aged 15-49 years with a live birth in the two years preceding the survey. Temporal changes in the use of caesarean section in each of the nine countries were calculated using generalised linear models and presented as risk differences(RD) with 95% confidence intervals(95%CI). Caesarean section was disaggregated against household wealth index and type of healthcare facility.

**Results:** Use of caesarean section ranged from 57.3% (95%CI:55.6–59.1%) in Egypt to 5.7% of births (95%CI:4.9–6.6%) in Yemen. Overall, the use of caesarean section has increased across the MENA region, except in Jordan, where there was no evidence of change (RD −2.3% (95%CI:-6.0–1.4%). Within all MENA countries, caesarean section use was highest in the richest quintile compared poorest quintile, for example, 43.8% (95%CI:38.0-47.6%) vs. 22.6% (95%CI:19.6-25.9%) in Iraq, respectively. Caesarean section was higher in private sector facilities compared to public sector: 21.8% (95%CI:18.2-25.9 %) vs. 15.7% (95%CI:13.3-18.4%) in Yemen, respectively.

**Conclusion:** Variations in caesarean section exist within and between Arab countries, and it was more commonly used amongst the richest quintiles and in private healthcare facilities. The private sector has a prominent role in the trends. Urgent policies and interventions are required to address non-medically indicated intervention.

## Introduction

Caesarean section is a lifesaving obstetric surgery that reduces maternal morbidity and mortality, (1) however both its under use and over use illustrate lack of appropriate care (2). Caesarean section usage has been increasing over the past decades, nonetheless, this rise is not mirrored by similar improvement in maternal and neonatal mortality (1, 3). Optimal caesarean section usage is believed to range between 10-15% to prevent maternal and neonatal mortality and morbidity, whilst rates above 20% have been shown not to improve maternal and neonatal mortality (1, 4, 5).

Although a vital part of obstetric medicine, caesarean section should be avoided unless clinically indicated due to the potential complications for mother and child (6). In particular, in the short term caesarean section increases the risk of haemorrhage, organ injury, infection and anaesthetic complications (7). In the longer term it is a leading risk factor for placental spectrum disorders and uterine rupture (8) and may have long term complications for infants including asthma and obesity(9, 10). The risk of these adverse outcomes in subsequent pregnancies increases with repeated caesarean section (11-13). Specifically, in resource-constrained settings, the risk of maternal deaths and the experiences of adverse outcomes such as a near miss is higher following a caesarean section compared to vaginal birth,(14) and the risk increases with repeated caesarean sections (15). In particular, a systematic review showed that maternal mortality following a caesarean section was 100 times higher in low resource settings compared to high income countries (16).

In parallel to the global overuse of caesarean sections, large inequalities in its use between different regions of the world and within countries have been reported, including the MENA region (17, 18). Within country inequalities in low and middle-income countries are represented by a higher use among the richest quintiles, among the more educated women and in private facilities (3, 17, 19). Considering the strong and growing influence of the private health care sector in the MENA region,(20) a closer look into inequalities can provide a better understanding of the healthcare systems in the region. This study aims to examine the trends of caesarean section and describe variations in caesarean section by economic status and type of healthcare facility (private/public sector).

## Methods

### Data

This was a secondary data analysis of the two most recent demographic and health surveys (DHS) or multiple indicator cluster surveys (MICS) for nine countries in the Middle East and North Africa (MENA) region, conducted between 2008 and 2020. These are publicly available data from (http://dhsprogram.com) or (https://mics.unicef.org/). DHS/MICS are nationally representative household surveys of women at reproductive ages, their infants and households, which are collected in low and middle-income countries. The sampling frame for each national survey includes area units across the entire country and the employed sampling procedure is a multi-stage stratified cluster sampling. Data are collected by trained interviewers using a standard questionnaire from each household. The questionnaire includes self-reported information about sociodemographic, household characteristics and health modules, including maternal and child health and details on delivery. Both the survey design and questionnaires are similar between MICS and DHS, across countries and over time, which allows for valid comparisons (21).

### Setting

All countries within the MENA region where Arabic is the national language and had available national level DHS/MICS data between 2008-2020 were included in the analysis. Countries with available data were: Algeria, Egypt, Iraq, Jordan, Palestine (State of Palestine or Palestinian territories), Qatar, Sudan, Tunisia and Yemen. Countries that had multiple surveys over the survey period were included for the trend analysis.

### Population

The sample population for this study included women aged 15-49 years who had a live birth in the two years preceding the survey date and those who reported on the mode of delivery for their last birth. Self-reported data on caesarean section were derived using the following questions for the DHS and MICS surveys respectively: “*Was (NAME) delivered by caesarean, that is, did they cut your belly open to take the baby out?*”. Self-reported measures of maternal indicators have been shown to be valid and reliable (22, 23). In a number of datasets, mode of delivery was missing for women that gave birth at home or outside of healthcare facilities; these women were recoded as having vaginal births. Women who did not answer the question on mode of delivery were excluded from the analysis; this proportion never exceeded 3.7% among eligible women. A small number of women had multiple births in the two years preceding the survey, and for the purposes of this study, the analysis only included data on the last live birth (one birth per woman).

### Variables

The primary covariates that were explored in the analysis were type of healthcare facility (Private, Public, home or other) and quintiles of wealth index. Public facilities were those defined by the MICS and DHS questionnaire, in general they included government-operated hospitals and clinics. Private sector facilities included private doctors, clinics or offices and hospitals, and facilities operated by other private organisations such as non-governmental organisations.(19). Wealth index is a measure of economic status and was constructed using ownership of household assets (24).

### Comparability and harmonisation

To ensure comparability across MICS and DHS questionnaires, women were included if they reported a live birth in the two years preceding the survey. MICS surveys ask women about births in the two years preceding the survey, whilst DHS asks women about births in the preceding five years. To reduce the recall period in the DHS dataset to match that of the MICS, we restricted the sample to women who delivered in the two years preceding the survey using the date of interview and date of last birth. Each data item in the DHS and MICS surveys were mapped for comparability and were recoded, if necessary, to create a harmonised variable.

### Data analysis

The prevalence estimates of caesarean section were calculated with 95% confidence intervals. The absolute annual change was calculated by subtracting the caesarean section rate in the latest survey from the previous survey, divided by the number of years between the two surveys. Trends over the two surveys were also calculated using generalised linear models and presented as an absolute risk differences between the two survey periods with 95% confidence intervals. Caesarean section rates were disaggregated by household wealth index and type of healthcare facility (private/public sector) and presented using equiplots and line graphs. The absolute number of births by healthcare facility type and the proportion of caesarean sections that occurred within the type of healthcare facility are presented using bar graphs. The complex survey design was accounted for in the analysis; in particular, proportions were weighted and accounted for clustering and stratification in the survey design. Analyses were conducted using STATA version 15. This was an unfunded study.

### Ethical Approval

This study was a secondary data analysis of publically available MICS and DHS data. As a result, ethical approval from the American University of Beirut was not required.

## Results

### Sample

The final sample size of the included surveys ranged from 13,986 women in Iraq’s 2011 survey, to 767 women in Qatar’s 2012 survey. The number of women included from each country and survey year is summarised in S1 Table. Three countries only had one survey available during the study period, and these are Algeria, Qatar and Yemen. Whilst Egypt, Jordan, Iraq, Palestine, Sudan and Tunisia had two surveys and were analysed as repeat cross sections. The Sudanese data did not include granular information on facility type in 2010, and the data from Qatar lacked the wealth index variables so these two countries were excluded from the respective disaggregated analyses.

### Caesarean section rates

In the most recent survey, use of caesarean section ranged from 5.7% of births (95%CI:4.9-6.6%) in Yemen to 57.3% of births (95%CI: 55.6–59.1%) in Egypt. Other than Sudan and Yemen, use of caesarean section was above 10% in all other countries (Figure 1). Overall, the proportion of caesarean sections out of all births increased between the first and second survey period in all MENA countries included in the analysis, except for Jordan. The largest increase occurred in Egypt, an average annual change of 4.5% or a risk difference of 26.7% (95%CI: 24.1-29.3%) between the periods covered by the two surveys (S2 Table). There were similarly large increases in Tunisia between 2010-2012 and 2016-2018 and Iraq between 2009-2011 and 2016-2018 with risk differences ranging from 11-17%. Jordan was an exception as it had no statistically significant change between 2010-2012 and 2016-2018 shown by a risk difference of −2.3 (95%CI: −6.0 - 1.4).

**Figure 1.**
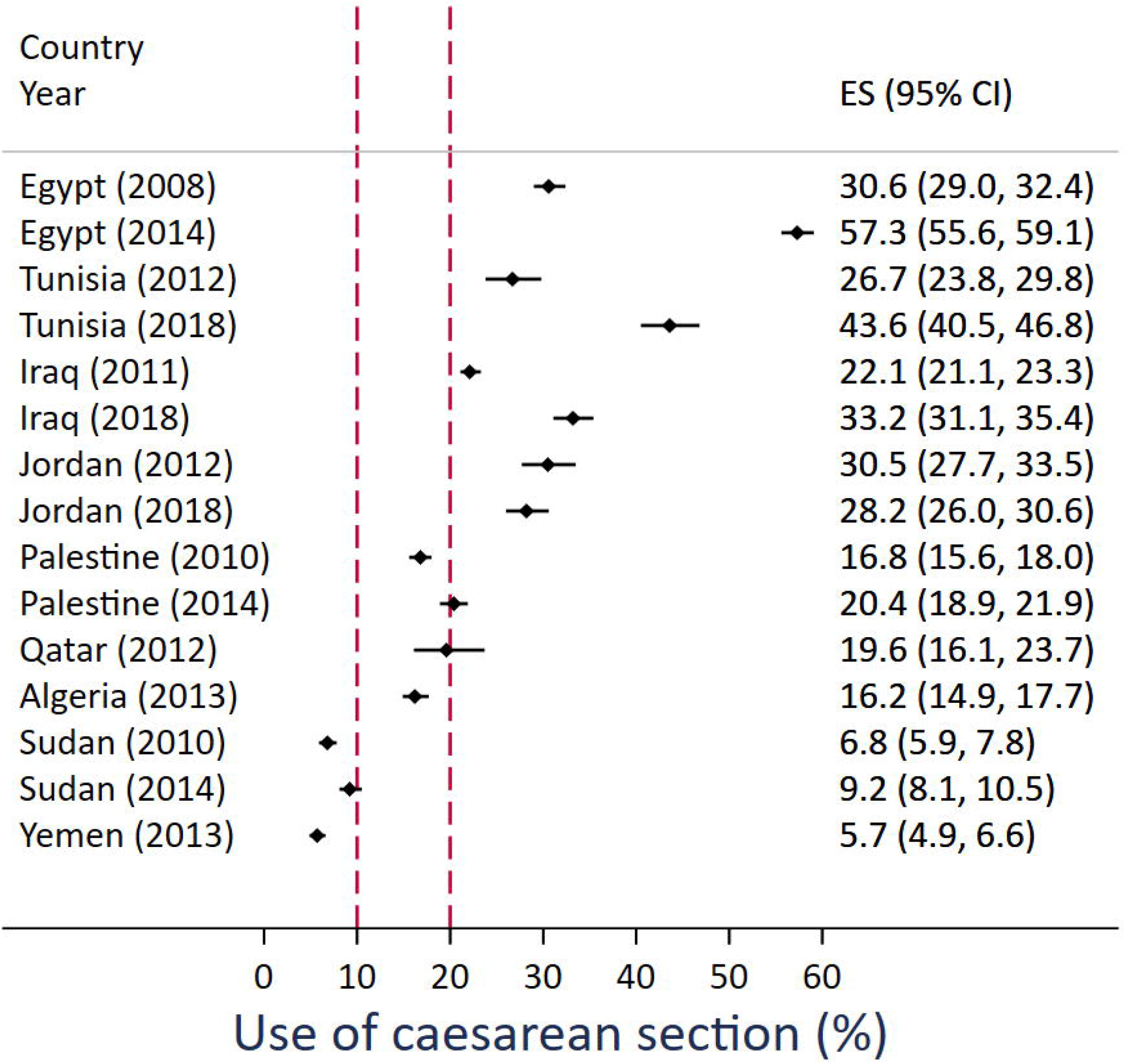
Proportion of caesarean sections at last birth that occurred in the previous two years before the survey in nine countries in the Middle East and North Africa. Footnote: Red line indicates underuse and overuse of caesarean section.

### Equity analysis

The use of caesarean section varied broadly across wealth quintiles for Egypt, Iraq, Sudan, Yemen and Tunisia (Figure 2 and S2 Table). For example, in the Egyptian 2014 survey, the use of caesarean section ranged from 43.4% (95% CI:39.8-47.1%) in the most deprived quintile to 72.4% (95% CI: 69.3-75.4%) in the wealthiest quintile. In Yemen, the proportion of caesareans was higher for the top wealth quintile reaching 14.6% (95% CI: 11.9-17.8%). In all countries, across all survey years, the highest use of caesarean section was among the wealthiest quintiles and the lowest use was among the most deprived quintiles; for example, in Iraq’s 2018 survey: 42.8% (95%CI:38.0–47.6%) vs. 22.6% (95%CI:19.6-25.9%), respectively. In countries that had multiple surveys, the use of caesarean section increased in all wealth quintiles between the two cross sections. Jordan was an exception and experienced a decline in the use of caesarean section in the periods covered by the 2012 and 2018 surveys in all quintiles other than the second most deprived quintile. In 2018, there was no difference in the caesarean section usage across wealth quintiles in Jordan.

**Figure 2.**
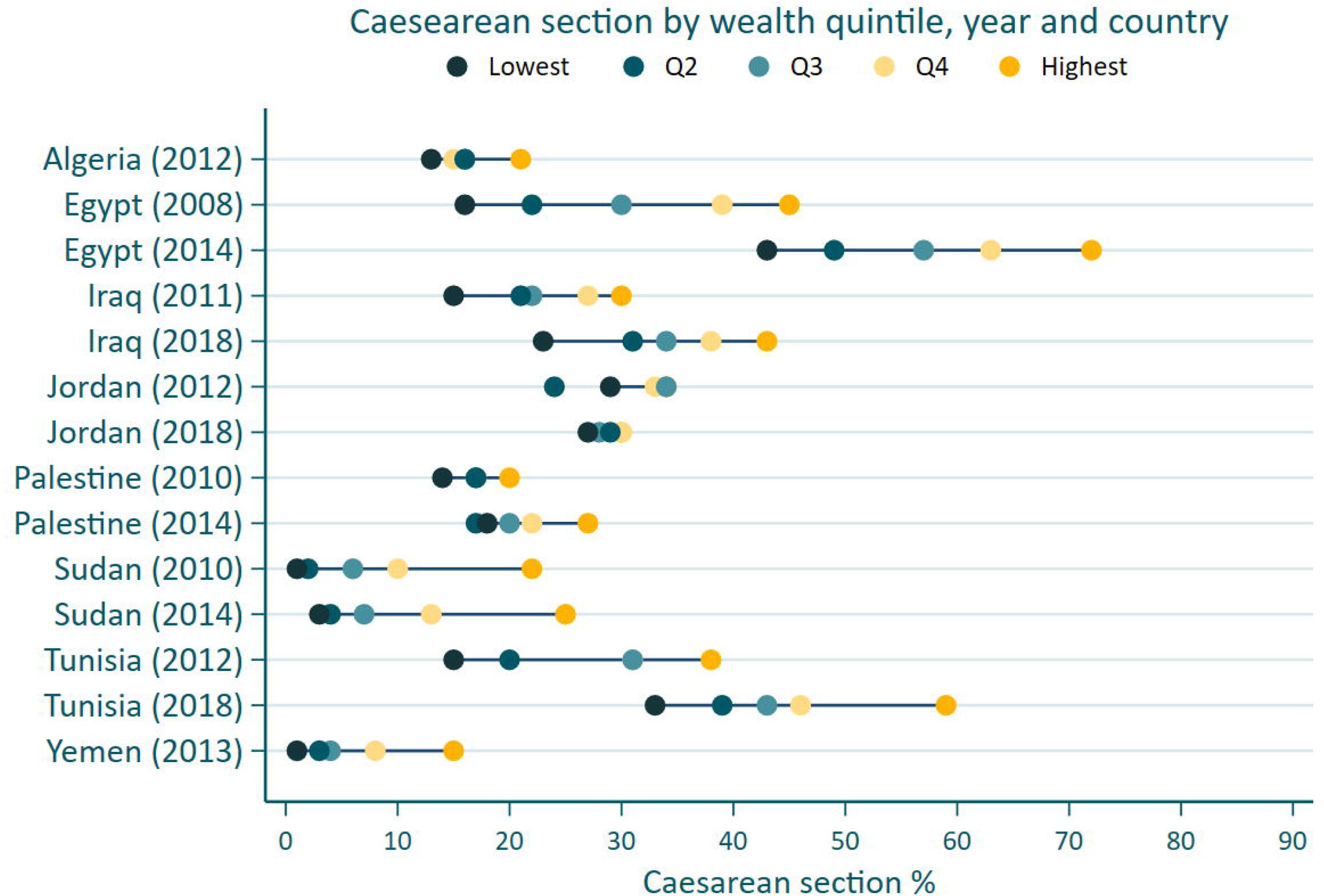
Proportion of caesarean section country, survey period and wealth quintile.

### Facility sector

Within the majority of countries included in the analysis, caesarean section use was highest in private sector facilities compared to public sector facilities; for example, 70.2% (95%CI:68.2-72.1%) vs. 50.9% (95%CI:47.6-54.1%) in Egypt 2012-2014, and 21.8% (95%CI:18.2-25.9 %) vs. 15.7% (95%CI:13.3-18.4%) in Yemen 2012-2014, respectively (Figure 3 and S2 Table). The use of caesarean section was similar in both private and public sector facilities in Palestine during both survey periods. In most countries that had multiple surveys, the use of caesarean section increased over time in both the private and public sector facilities, other than Jordan where it declined from 29.6% (95%CI: 26.9-32.5%) to 26.2% (95%CI: 23.9-28.7%) in the public sector between the two survey periods. In the majority of countries, the absolute proportion of births was highest in public sector facilities, other than Egypt, where more births occurred in the private sector (Figure 4). The proportion of all births (both vaginal and caesarean section) in the private sector increased in Egypt, Iraq and Tunisia between survey periods, while it remained unchanged in Jordan.

**Figure 3.**
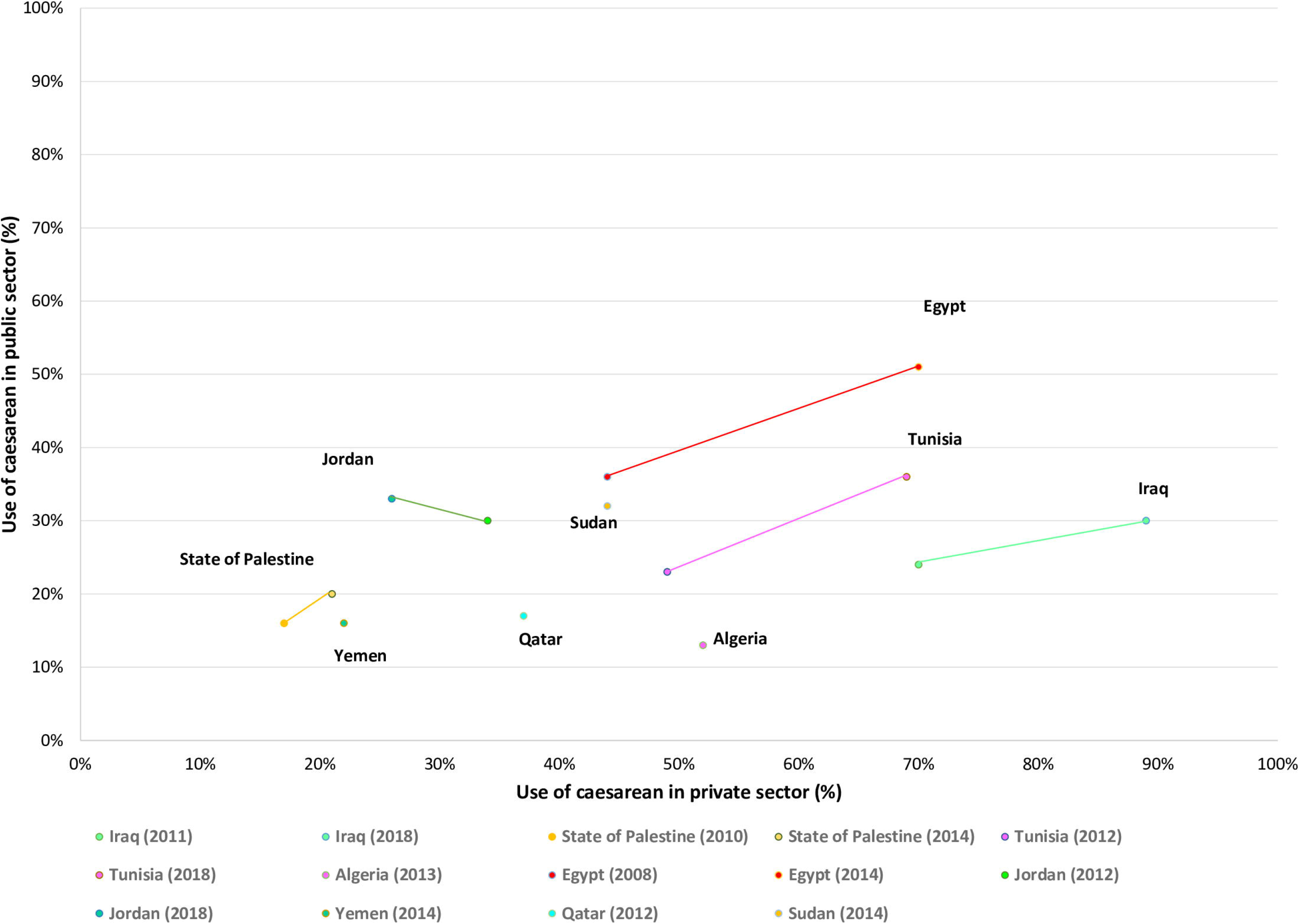
The use of caesarean section in private and public sectors by country and survey period.

**Figure 4.**
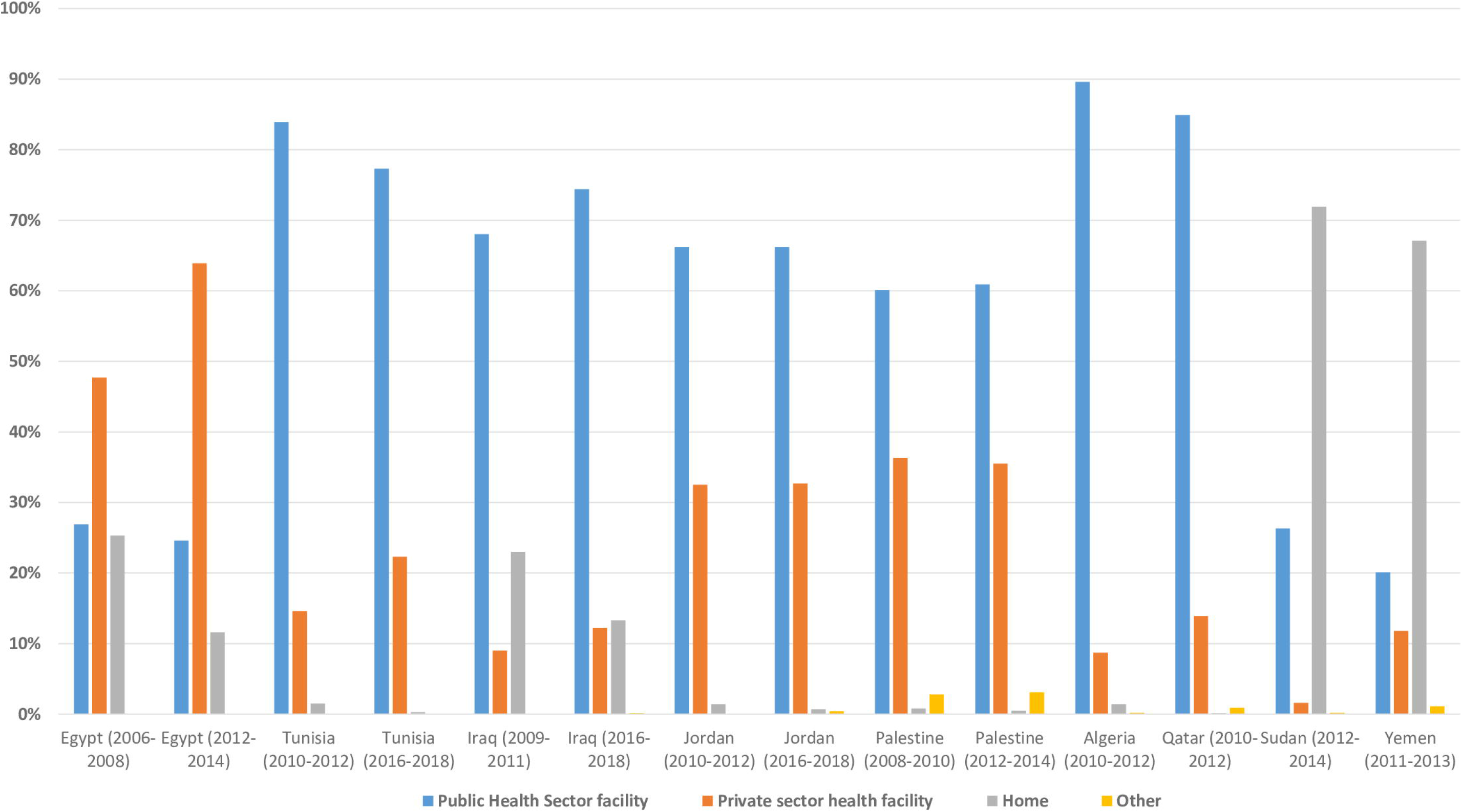
Proportion of all births according to place of delivery for the last two surveys.

The distribution of caesarean section by wealth quintile amongst private and public sector facilities is presented in figure 5. In general, within the private sector, the highest use of caesarean section was in the wealthiest quintiles. Sudan, Iraq, Tunisia and Algeria were exceptions where the highest use of caesarean section was in the middle and lowest quintiles, respectively. In the public sector, use of caesarean section followed a similar trend of highest use in the wealthiest quintile and lowest in the most deprived quintiles for Palestine, Yemen, Tunisia, and Egypt. In the private sector, there was a broad inequality between the poorest and wealthiest quintiles, while in the public sector, the range between the poorest and least wealth quintiles was smaller.

**Figure 5.**
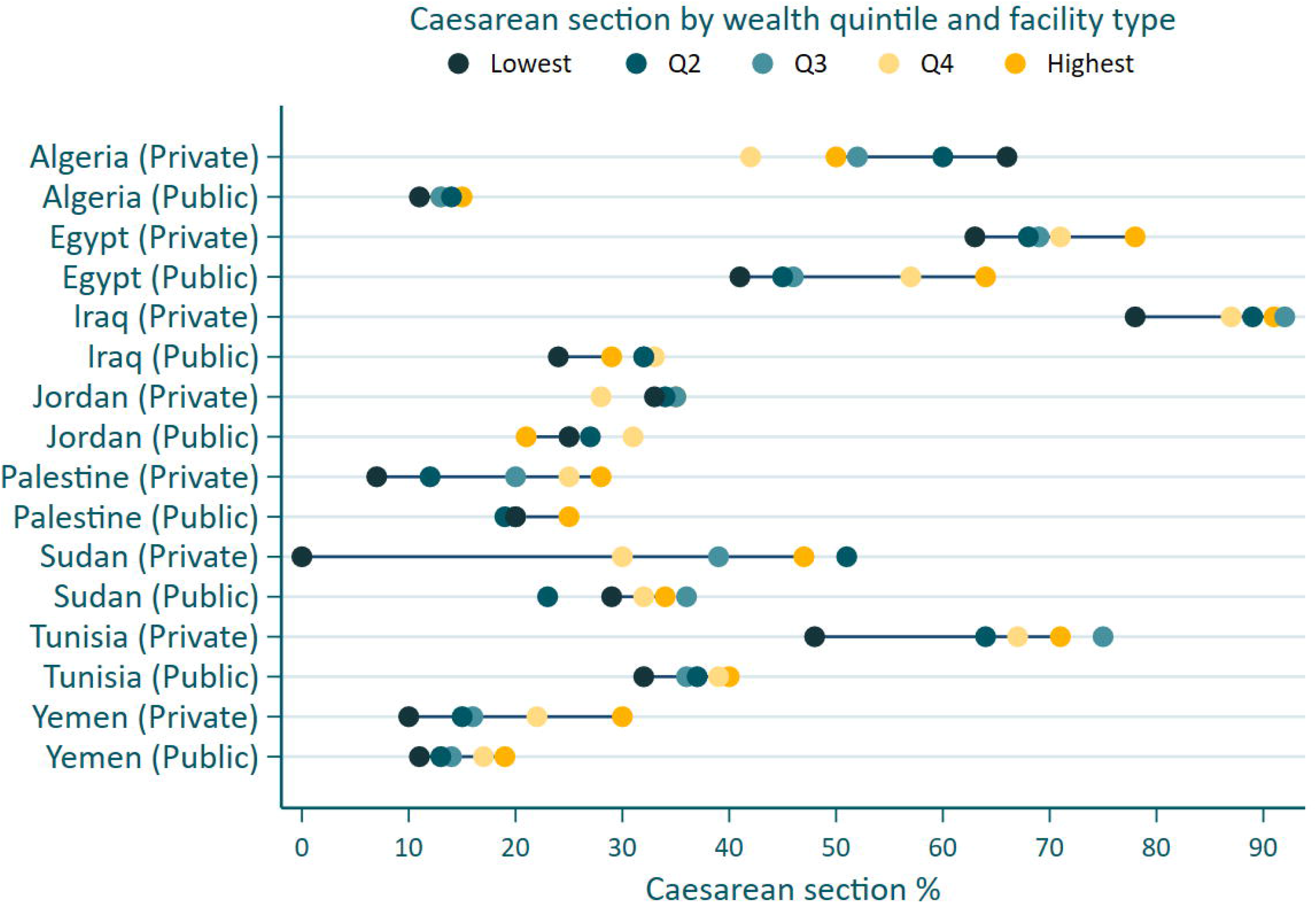
Caesarean section delivered in public and private sectors, by wealth quintile and country, for the most recent survey. Footnote: Last DHS/MICS survey was used. Qatar did not collect wealth quintile.

## Discussion

### Main findings

There is large disparity in the use of caesarean section amongst the nine countries of the MENA region. Notably, there was a high use of caesarean section in Egypt, Tunisia and Iraq whilst it was low and likely inadequate in Yemen and Sudan. Caesarean section use increased across time in all countries, excluding Jordan, which had no change between the survey periods. In general, the highest usage of caesarean section was in the private sector and among the wealthiest populations. Over time, caesarean section use increased in both the public and private sector facilities with an increase in private sector births in many countries. For the most part, the majority of births occur in the public sector in Arab countries of the MENA region, modest increases in caesarean section use in the public sector will result in larger absolute increases in the use of caesarean section overall.

### Strengths and limitations

This study was limited to countries that had an available MICS or DHS surveys, there were lack of data from many gulf countries, whilst nearly all MENA nations in North Africa had available data. The gulf countries represent the wealthiest Arab nations and have more advanced health systems. In addition, data that was collected through household surveys, and therefore lacked granularity in terms of obstetric history and whether the caesarean section was clinically indicated. In particular, utilising the Robson criteria in future studies based on hospital data would help us understand the proportion of caesarean sections that could have been avoided. Despite these limitations, this study is comprehensive in terms of its exploration of the role of the private sector and relative wealth in the use of caesarean section in the MENA region.

### Interpretation

In comparison to other regions, the MENA region has some of the highest use of caesarean section, globally (3, 17). The results are consistent with the global studies that have shown an increasing trend in caesarean section use during the last decade (3). In particular, Egypt has one of the highest usage of caesarean section in the world (3) and this study builds on previous literature to show that the rate of caesarean section is increasing across time (25). In Arab countries midwifery models of obstetric care are rarely implemented at a national level due to a shortage of midwives;(26) this is likely to be a contributing factor to the large proportion of caesarean sections in the region (27).

There were also large disparities within the MENA region, as shown by the low usage in Yemen and Sudan, which may be explained by a context of limited resources, fragile healthcare systems and conflict. Yemen has a very low facility birth rate with approximately 70% of births occurring at home (28). The country’s DHS survey report highlighted that there were 26 neonatal deaths and 48 maternal deaths per 100,000 live births with 42% of women dying at home. Similarly, the maternal mortality ratio across Sudanese regions ranges from 63 to 260 maternal deaths per 100,00 live births (29). In other words, there were likely women in Yemen and Sudan who required lifesaving care in the form of a caesarean section but did not receive the intervention.

Similar to previous studies completed in the region: Jordan, Iraq and Egypt had higher caesarean section births in private hospitals compared to public hospitals (25, 30). This relationship is likely driven by a multitude of factors including financial gain, fear of litigation and time convenience for healthcare providers (18, 31, 32) as well as women’s fear of vaginal birth and labor pain,(32-34) lack of choices in models of care for women and the over-medicalisation of childbirth (35-38). Previous studies completed in the region have indicated that growth in the private sector is a key driver to the increase in caesarean section rate (25). Many countries in this study had a background increase in the use of private hospitals in all births, which further suggests that the private sector has a pivotal role in increasing caesarean section usage in the region. The rise of the private sector is multifactorial as many countries do not have national health insurance program, private insurance is incentivised for the rich, and public perception of private care is that it is better due ‘on demand’ access and the availability of the latest medical technologies (39). However, the private sector is often poorly regulated and private hospitals are often run with near complete autonomy from the public health system (31, 40, 41).

A previous study in Gaza, Palestine, showed that caesarean section was more commonly performed in governmental facilities, while there was no association between facility type and caesarean section in the West Bank. This study examined the country level proportions in Palestine and found similar proportions of caesarean births in both private and public hospitals (42). It is possible that regional level differences in the use of caesarean section between facilities exists. The contextual factors of rising poverty has made the private sector inaccessible to much of the population; in addition, governmental hospital services are available in all areas and Government Health Insurance, which includes maternity health service, is available at little cost (43-45).

Given the relationship between caesarean section use and the role of the private sector, it is no surprise that the highest usage is in the wealthiest and most affluent populations. In addition, the private sector plays a critical role in the extension of inequalities in caesarean section use in the MENA. The inequality gap between the wealthiest and poorest shown in this study is consistent with previous studies in other low and middle income countries (17, 46-48).

It is interesting to note that there was no change over time in caesarean section usage in Jordan and the inequality gap in the usage of caesarean section did not exist in the most recent survey. The findings are in contrast with previous reports of a continuous increasing trend in cesarean section use in Jordan between 2009 and 2012 (25). Future studies should aim to understand the mechanisms for the decrease in the inequality gap in caesarean section, reduced role of the private sector and stability of the usage of caesarean section in Jordan.

In general, it is likely large proportions of caesarean sections in the MENA region were not medically indicated; thus the implementation of non-clinical interventions may reduce unnecessary caesareans sections (49). Betran et al. proposed that structural interventions should target drivers of high use of caesarean section, recognise the context, and be adapted to women’s views, cultural norms and clinical practice at the individual and structural levels (49).

## Conclusion

Variations in the use of caesarean section exist within and between Arab countries in the MENA region, where over-use of interventions is an established routine in some contexts and in others with limited resources, life-saving interventions are unavailable when needed. The MENA region is in need of optimizing the use of caesarean section that includes ensuring access to safe caesarean section when it is required coupled with health system reform and non-medical interventions to address non-medically indicated caesarean sections.

## Data Availability

Data are available at: https://mics.unicef.org/ & https://dhsprogram.com/data/

https://dhsprogram.com/data/

## Significance

A number of Arab countries such as Egypt and Lebanon have known high use of caesarean section; however, more recent data is required to confirm if use of caesarean section is uniform, increasing and related to the private sector in Arab countries. Both extremes of caesarean section use were present in the Arab region. Use of caesareans section has increased over time; however, there was no change in Jordan. The largest proportion of caesareans occur in the private sector; however, an increase in caesarean sections in the public hospitals is concerning as the majority of births occur in this sector. There is a requirement to act to reduce non-necessary caesareans sections in the Arab region.

## Availability of data and material

Data are available at: https://mics.unicef.org/ & https://dhsprogram.com/data/

## Author contributions

SM and TK conceptualized the study. SM led the analysis, project management and wrote the first draft. NA, AS and CO provided guidance on the analysis and drafted the edited the manuscript. TK and MA interpreted the data and drafted the manuscript. SM is the guarantor.

## Acknowledgements

No acknowledgements

## Supporting information

S1 Table. Sample selection

S2 Table. The change in the proportion of caesarean section over time

