## Supplementary tables for "Trends, wealth inequalities and the role of the private sector in caesarean section in the Middle East and North Africa: a repeat cross-sectional analysis of population-based surveys"

S1 Table. Sample selection

| **Country** | **MICS** | **DHS** | **Study**  **period** | **Original Sample size** | **No consent/ Not completed** | **Never had a child** | **No births in the last 2 years** | **Eligible women** | **Excluded:**  **delivery mode missing** | **Final study sample** |
| --- | --- | --- | --- | --- | --- | --- | --- | --- | --- | --- |
| **Iraq** | 2011 |  | 2009-2011 | 56,445 | 1,251 | 4,079 | 37,121 | 13,994 | 8 | 13,986 |
|  | 2018 |  | 2016-2018 | 31,060 | 400 | 2,254 | 22,156 | 6,250 | 2 | 6,248 |
| **Palestine** | 2010 |  | 2008-2010 | 12,322 | 343 | 845 | 6,805 | 4,329 | 2 | 4,327 |
|  | 2014 |  | 2011-2014 | 13,964 | 597 | 732 | 9,744 | 2,891 | 5 | 2,886 |
| **Tunisia** | 2012 |  | 2009-2012 | 10,514 | 299 | 541 | 8,539 | 1,135 | 1 | 1,134 |
|  | 2018 |  | 2016-2018 | 11,017 | 458 | 621 | 8,726 | 1,212 | 1 | 1,211 |
| **Qatar** | 2012 |  | 2010-2012 | 5,809 | 110 | 2734 | 2,195 | 770 | 3 | 767 |
| **Algeria** | 2013 |  | 2010-2013 | 41,184 | 2,637 | 2,706 | 29,863 | 5,978 | 111 | 5,867 |
| **Sudan** | 2010 |  | 2008-2010 | 18,614 | 1,440 | 6,334 | 5,213 | 5,627 | 32 | 5,595 |
|  | 2014 |  | 2012-2014 | 20,327 | 2,025 | 6,601 | 6,024 | 5,677 | 60 | 5,617 |
| **Egypt** |  | 2008 | 2006-2008 | 16,527 | 0 | 1,749 | 10,243 | 4,535 | 3 | 4,532 |
|  |  | 2014 | 2012-2014 | 21,762 | 0 | 1992 | 13,354 | 6,416 | 0 | 6416 |
| **Jordan** |  | 2012 | 2010-2012 | 11,352 | 0 | 1,048 | 6,626 | 3,678 | 58 | 3620 |
|  |  | 2018 | 2016-2018 | 14,689 | 0 | 1,774 | 9,006 | 3,909 | 145 | 3,764 |
| **Yemen** |  | 2013 | 2011-2013 | 16,656 | 0 | 1,968 | 8,515 | 6,173 | 11 | 6,162 |

S2 Table.. The change in the proportion of caesarean section over time.

| **Country** | **Period 1 (2008-2012)** | **Period 2 (2013-2018)** | **Average yearly change** | **Absolute risk difference (95% CI)** |
| --- | --- | --- | --- | --- |
| **Egypt** | 30.6 (29.0; 32.4) | 57.3 (55.6; 59.1) | 4.45% | 26.7 (24.1-29.3) |
| **Tunisia** | 26.7 (23.8; 29.8) | 43.6 (40.5; 46.8) | 2.82% | 16.9 (12.6-21.3) |
| **Iraq** | 22.2 (21.1; 23.3) | 33.2 (31.1; 35.4) | 1.59% | 11.0 (8.6-13.4) |
| **Jordan** | 30.5 (27.7; 33.5) | 28.2 (26.0; 30.6) | -0.38% | -2.3 (-6.0 - 1.4) |
| **Palestine** | 16.8 (15.6; 18.0) | 20.4 (18.9; 21.9) | 0.9% | 3.6 (1.6-5.6) |
| **Sudan** | 6.8 (5.9; 7.8) | 9.2 (8.1; 10.5) | 0.6% | 2.4 (0.9-4.0) |
| **Qatar** |  | 19.6 (16.1; 23.7) | -- | -- |
| **Algeria** |  | 16.2 (14.9; 17.7) | -- | -- |
| **Yemen** |  | 5.7 (4.9; 6.6) | -- | -- |

Values are percentages (95% Confidence intervals).

Supplementary Table 3. Use of caesarean section by wealth index and delivery place by country and survey year (Proportion and 95% CI)

|  | **Wealth quintiles** | | | | | | | | | | **Facility type** | | | |
| --- | --- | --- | --- | --- | --- | --- | --- | --- | --- | --- | --- | --- | --- | --- |
|  | **Poorest** | **95 % CI** | **Second** | **95 % CI** | **Middle** | **95 % CI** | **Fourth** | **95 % CI** | **Richest** | **95% CI** | **Public** | **95 % CI** | **Private** | **95 % CI** |
| **Iraq** |  |  |  |  |  |  |  |  |  |  |  |  |  |  |
| **2011** | 14.6% | 13; 16.2 | 20.5% | 18.4; 22.8 | 21.7% | 19.4; 24.2 | 27.4% | 24.5; 30.4 | 30.0% | 26.8; 33.4 | 23.7 | 22.4; 25.1 | 70.4 | 64.9; 75.3 |
| **2018** | 22.6% | 19.6; 25.9 | 30.6% | 25.6; 36.1 | 33.8% | 30.2; 37.6 | 38.4% | 34.2; 42.8 | 42.8% | 38; 47.6 | 30.1 | 27.8; 32.5 | 88.9 | 84.3; 92.3 |
| **State of Palestine** |  |  |  |  |  |  |  |  |  |  |  |  |  |  |
| **2010** | 13.7% | 11.5; 16.2 | 17.0% | 14.8; 19.4 | 17.4% | 14.9; 20.2 | 17.3% | 14.7; 20.3 | 19.7% | 16.4; 23.3 | 16.4 | 15.1; 17.9 | 17.4 | 15.4; 20 |
| **2014** | 17.7% | 14.8; 20.9 | 17.1% | 14.3; 20.3 | 20.4% | 17.3; 23.8 | 21.9% | 18.8; 25.4 | 26.7% | 22.7; 31.2 | 20.3 | 18.5; 22.2 | 20.8 | 18.4; 23.4 |
| **Tunisia** |  |  |  |  |  |  |  |  |  |  |  |  |  |  |
| **2012** | 15.3% | 10.9; 20.9 | 19.7% | 14.2; 26.7 | 31.1% | 24.1; 39.2 | 30.7% | 24.5; 37.8 | 37.7% | 29.8; 46.3 | 23.3 | 20.1; 26.8 | 49.2 | 39.9; 58.6 |
| **2018** | 32.7% | 27.1; 38.9 | 39.0% | 32.7; 45.7 | 43.5% | 36.5; 50.7 | 46.1% | 39.9; 52.3 | 59.6% | 50.3; 66.4 | 36.5 | 33.2; 39.9 | 69 | 62.4; 74.9 |
| **Qatar** |  |  |  |  |  |  |  |  |  |  |  |  |  |  |
| **2012** |  |  |  |  |  |  |  |  |  |  | 17.1 | 13.5; 21.3 | 37 | 24.1; 52 |
| **Algeria** |  |  |  |  |  |  |  |  |  |  |  |  |  |  |
| **2013** | 13.4% | 10.9; 16.4 | 16.2% | 13.4; 19.4 | 16.1% | 13.4; 19.2 | 15.3% | 12.6; 18.5 | 21.3% | 18; 25 | 13 | 11.7; 14.4 | 52.1 | 45.6; 58.5 |
| **Egypt** |  |  |  |  |  |  |  |  |  |  |  |  |  |  |
| **2008** | 16.2% | 13.8; 19 | 22.4% | 19.3; 25.8 | 29.9% | 26.6; 33.4 | 39.1% | 35.5; 42.8 | 45.5% | 41.5; 49.4 | 36 | 32.8; 39.2 | 44 | 41.5; 46.5 |
| **2014** | 43.4% | 39.8; 47.1 | 49.3% | 45.8; 52.8 | 57.4% | 54.2; 60.6 | 63.3% | 60; 66.4 | 72.4% | 69.3; 75.4 | 50.9 | 47.6; 54.1 | 70.2 | 68.2; 72.1 |
| **Jordan** |  |  |  |  |  |  |  |  |  |  |  |  |  |  |
| **2012** | 29.3% | 24.5; 34.7 | 24.3% | 20.4; 28.7 | 33.7% | 28.2; 39.5 | 33.3% | 26.5; 40.9 | 33.5% | 25; 43.2 | 29.6 | 26.9; 32.5 | 33.7 | 28.2; 39.7 |
| **2018** | 26.5% | 23; 30.3 | 28.5% | 24.4; 33.1 | 27.8% | 23; 33.1 | 30.1% | 24.4; 36.6 | 29.9% | 20.8; 30.6 | 26.2 | 23.9; 28.7 | 32.9 | 28.5; 37.6 |
| **Sudan** |  |  |  |  |  |  |  |  |  |  |  |  |  |  |
| **2010** | 1.0% | 0.5; 1.9 | 2.0% | 1.3; 3.1 | 5.8% | 4.4; 7.6 | 9.7% | 7.4; 12.7 | 22.0% | 18.4; 26 |  |  |  |  |
| **2014** | 2.6% | 1.6; 4 | 3.7% | 2.6; 5.1 | 7.0% | 5.1; 9.6 | 12.8% | 9.8; 16.5 | 25.3% | 21.5; 29.4 | 32.1 | 28.6; 36 | 44.3 | 31; 58.4 |
| **Yemen** |  |  |  |  |  |  |  |  |  |  |  |  |  |  |
| **2014** | 1.4% | 0.8; 2.3 | 2.6% | 1.8; 3.7 | 4.4% | 3.3; 5.9 | 8.3% | 6.4; 10.6 | 14.6% | 11.9; 17.8 | 15.7 | 13.3; 18.4 | 21.8 | 18.2; 25.9 |
